# Ultra-rare and common genetic variant analysis converge to implicate negative selection and neuronal processes in the aetiology of schizophrenia

**DOI:** 10.1101/2021.05.26.21257794

**Authors:** Wonuola A Akingbuwa, Anke R Hammerschlag, Meike Bartels, Michel G Nivard, Christel M Middeldorp

**Author notes:** **Corresponding Author:** Wonuola Akingbuwa, MSc, Department of Biological Psychology, Vrije Universiteit Amsterdam, Van der Boechorststraat 7, 1081 BT Amsterdam, the Netherlands. Jointly supervised the work (Michel G Nivard, Christel M Middeldorp).

## Abstract

Both common and rare genetic variants (minor allele frequency > 1% and < 0.1% respectively) have been implicated in the aetiology of schizophrenia. In this study, we integrate single-cell gene expression data with publicly available Genome-Wide Association Study (GWAS) and exome sequenced data in order to investigate in parallel, the enrichment of common and (ultra-)rare variants related to schizophrenia in several functionally relevant gene-sets. Four types of gene-sets were constructed 1) protein-truncating variant (PTV)-intolerant (PI) genes 2) genes expressed in brain cell types and neurons ascertained from mouse and human brain tissue 3) genes defined by synaptic function and location and 4) intersection genes, i.e., PI genes that are expressed in the human and mouse brain cell gene-sets. We show that common as well as ultra-rare schizophrenia-associated variants are overrepresented in PI genes, in excitatory neurons from the prefrontal cortex and hippocampus, medium spiny neurons, and genes enriched for synaptic processes. We also observed stronger enrichment in the intersection genes. Our findings suggest that across the allele frequency spectrum, genes and genetic variants likely to be under stringent selection, and those expressed in particular brain cell types, are involved in the same biological pathways influencing the risk for schizophrenia.

## INTRODUCTION

Schizophrenia is severe and highly heritable, with onset in adolescence or early adulthood. It is associated with early mortality and reduced fertility^1, 2^, which put selective pressure on genetic variants related to schizophrenia. Despite apparent negative selection, schizophrenia remains highly heritable and relatively prevalent^3^.

There is evidence that both common (minor allele frequency (MAF) > 1%) and rare genetic variants are related to schizophrenia^4-6^, with an inverse relationship between variant effects and their frequency. Rarer variants can have larger effect, while common variants are maintained via background selection^7^, and can only persist in the presence of negative selection if their effects are small^8, 9^. However, common variants, despite smaller individual effects, collectively explain a substantial proportion of the genetic variance in schizophrenia^6, 10^. The presence of negative selection results in extreme polygenicity where substantial portions of the genome carry variants with tiny individual effects, yet only genes with critical effects when mutated would strongly impact the disorder^11^.

The genetic architecture of schizophrenia has forced two separate lines of enquiry into its aetiology. Genome wide associations studies (GWAS) have successfully targeted common variants,^6, 7, 12^ resulting in valuable leads for functional follow-up studies of individual loci. For example, in-depth study of the lead genome wide hit for SCZ in the Major Histocompatibility Complex (MHC) locus has implicated complement component 4 (C4) gene expression and possibly synaptic pruning in puberty in the aetiology of schizophrenia^13^. While further analysis of the schizophrenia locus in the *SLC39A8* gene implicated manganese (Mn) related brain phenotypes in the aetiology of schizophrenia^14^.

Whole exome sequencing (WES) has simultaneously been used to identify rare mutations with larger effects. To reduce the multiple testing burden, research has focused on specific classes of rare variants for which a deleterious (risk-increasing) effect can be assumed a-priori. Specifically, modest samples sizes have been leveraged by focusing on *singleton* (i.e. only observed once) variants in genes intolerant to mutations, that are predicted to be protein-truncating (PTVs) (i.e. disruptive and likely lead to loss of gene function). These variants and genes are the ones most likely to increase the risk of schizophrenia when perturbed by a mutation with a critical/negative effect^4, 15-17^. A recent study validated this analytical strategy and identified 10 genes in which ultra-rare deleterious variants were associated with schizophrenia^4^.

These parallel lines of common and rare variant-based genetic inquiry share a common goal: increased understanding of the neuro-biology of schizophrenia. Subsequently, functional analyses are crucial for understanding the pathways and mechanisms via which disorder-associated genetic variants may act. Functional analyses of common and rare variants detected in GWAS or WES studies have implicated the brain and brain-expressed genes in the aetiology of schizophrenia^6, 7, 15, 16^. More recently, high throughput single-cell RNA sequencing techniques, which are able to provide expression profiles of individual brain cells, allow us to associate traits with specific brain cell types. Single-cell expression data of mouse and human brain cells have revealed that schizophrenia-associated common and rare variants are enriched in genes expressed in (excitatory) neurons more than in other (non-neuronal) brain cells^4, 18, 19^.

In this preregistered study (https://osf.io/uyv2s), we integrated single-cell gene expression data with results from GWAS and exome sequenced data to investigate, in parallel, the enrichment of common and (ultra-)rare variants related to schizophrenia in specific brain cell types. We investigated whether trait-associated common and (ultra-)rare variants were enriched in sets of genes expressed in different brain cell types and neurons, as well as PTV-intolerant (PI) genes, which are under stringent selection. As synaptic functions have previously been implicated in the aetiology of schizophrenia, we included gene-sets based on synaptic processes and composition. Finally, we investigated gene-sets made up of the intersection of PI genes and brain-expressed genes (i.e. PI genes that are expressed in the brain) as these are potentially of considerable value for follow-up analyses if relevant. By synchronizing the functional analyses across common and rare variants, the current study attempts to answer two questions: 1) Do common and (ultra-)rare variant gene-set and cell type enrichment analyses converge to similar results for schizophrenia and if so, 2) What gene-sets are implicated across both (ultra-)rare and common variant analyses? The current analyses may shed light on whether common and (ultra-)rare variants reveal unique aspects of the aetiology of schizophrenia, or implicate the same pathways.

## METHODS

### Data and sources

#### GWAS data

Summary data of GWAS results of schizophrenia in individuals of European^7^ and East Asian ancestry^10^ were obtained. The European dataset included 105,318 individuals (40,675 cases and 64,643 controls) from 61 samples, while the East Asian dataset included 58,140 individuals (22,778 cases and 35,362 controls) from 16 samples.

#### Exome sequenced data

Genotype and phenotype data were obtained from the Swedish Schizophrenia Exome Sequencing Project^15^, a case-control sample of 12,380 unrelated Swedish individuals. Cases primarily had diagnoses of schizophrenia, although a small proportion of individuals were diagnosed with bipolar disorder.

#### Gene-sets

We selected genes with pLI > 0.9 from the Genome Aggregation Database (gnomAD) as protein-truncating variant (PTV)-intolerant (PI) genes, producing a list of 3063 genes^20^.

Human brain cell gene-sets were based on single-nucleus RNA-sequence (snRNA-seq) data generated on the Genotype-Tissue Expression (GTEx) project brain tissues^21^. We included a total of 14 cell types as ascertained in the study referenced. Excluding sporadic genes and genes with low expression, for the 14 cell types we selected the top 1600 (roughly 15%) differentially expressed genes in each cell type, which likely most of the genes that have specific functions in specific cell types.

Mouse brain cell gene-sets were based on data obtained from a previous study^19^. In that study, cells were assigned to level 1 classification, with subtypes of level 1 assigned as level 2 on the basis of single-cell RNA-sequence (scRNA-seq) data and clustering analyses. We focused our analyses on the 24 level 1 gene-sets. As the scRNA-seq data were from mouse brains, we mapped the gene homologs using the human-mouse homolog reference from Mouse Genome Informatics. We again selected the top 1600 differentially expressed genes in each cell type.

Synaptic gene-sets were selected based on synaptic gene ontology from the SynGo database^22^, including gene-sets defined by cellular component; the location in which the genes are active, or by biological process; the synaptic processes/functions they influence. In order to ensure that gene-sets were well powered, we selected only gene-sets containing 50 or more genes, resulting in a total of 35 gene-sets.

PI x brain cell gene-sets contained the intersection genes that are PTV-intolerant and are present in each human and mouse brain cell gene-set. Although the PI x brain cell type are smaller than either the PI-gene-set or the brain cell type specific gene-set, they are potentially more strongly enriched given the genes are (1) relevant to brain function and (2) under strong negative selection.

All 112 gene-sets analysed are available in Supplementary Table 1.

#### Common variant analyses

Common variant enrichment was evaluated using competitive analyses in MAGMA (v1.08b)^23^. MAGMA is a program for gene and gene-set analysis based on linear regression. First, gene-level associations are computed in which *p*-values for individuals SNPs are averaged taking linkage disequilibrium (LD) structure into account. We used the European panel of 1000 Genomes Project phase 3 for LD estimation in the GWAS based on individuals of European ancestry, and the East Asian panel in the GWAS based on individuals of East Asian ancestry. Subsequently, gene-based *p*-values were converted to z scores to test associations between each gene-set and schizophrenia diagnosis. For each GWAS summary dataset, we excluded duplicate SNPs, and SNPs with INFO <0.8. Gene location information with start and stop sites were obtained from the MAGMA website, with no windows specified around the genes.

Competitive analyses test whether enrichment in genes in a gene-set is greater than genes outside the gene-set. We thus tested enrichment of schizophrenia-associated common variants in our gene-sets of interest versus all other genes. Additionally, to determine whether the brain gene-sets were enriched for schizophrenia-associated common variants above other brain-expressed genes, we conditioned all brain and intersection gene-sets on only brain-expressed genes. This was done for all brain expressed genes, as well as the top 50%, 20% and 10% expressed genes. Construction of these gene-sets are described in the Supplementary text and the gene-sets are available in Supplementary Table 1.

Finally, we evaluated significant enrichment of the intersection gene-sets conditioned on their corresponding non-intersection brain gene-set, the PI gene-set, and both^24^. Significant associations in the intersection gene-sets after conditioning on brain-expressed genes suggest that the association is not due to association of genes expressed in the brain, significant associations after conditioning on PI genes suggest that the association is not due to association of PI genes, while significant associations after conditioning on both suggest that the gene-set is associated because of the combination of the brain expression and PI.

#### Rare variant analyses

Analyses of exome sequenced data including QC were performed using Hail 0.2 (Hail Team. Hail 0.2.62-84fa81b9ea3d. https://github.com/hail-is/hail/commit/84fa81b9ea3d). QC generally followed those described in a previous study^16^, and those detailed here: https://astheeggeggs.github.io/BipEx/index.html. Full description, including variant annotation and variant definitions are provided in the Supplementary text.

##### Gene-set (association) analyses

We assessed (ultra-)rare variant enrichment in each gene-set using logistic regression of schizophrenia diagnosis on (ultra-)rare variants burden scores. Additionally, we evaluated significant enrichment in the intersection gene-sets above the non-intersection brain gene-sets by including burden scores from non-intersection brain gene-sets, the PI gene-set, or both sets, in a single regression. Sex and 10 genetic PCs were included as covariates in all analyses. We excluded individuals who were more than 4 median absolute deviations from the study specific median number of synonymous (ultra-)rare variants. The significance threshold was set to 5% false discovery rate in analyses across all gene-sets per variant allele frequency. To ensure that population stratification within Sweden did not affect results, we created more genetically homogeneous subsets of the data, excluding predicted Northern-Swedish, Finnish and “other European” outliers from the analyses. We compared association results in the homogenous subsets to the total sample. This procedure and results are described in the Supplementary text and Figures 1-3.

**Figure 1:**
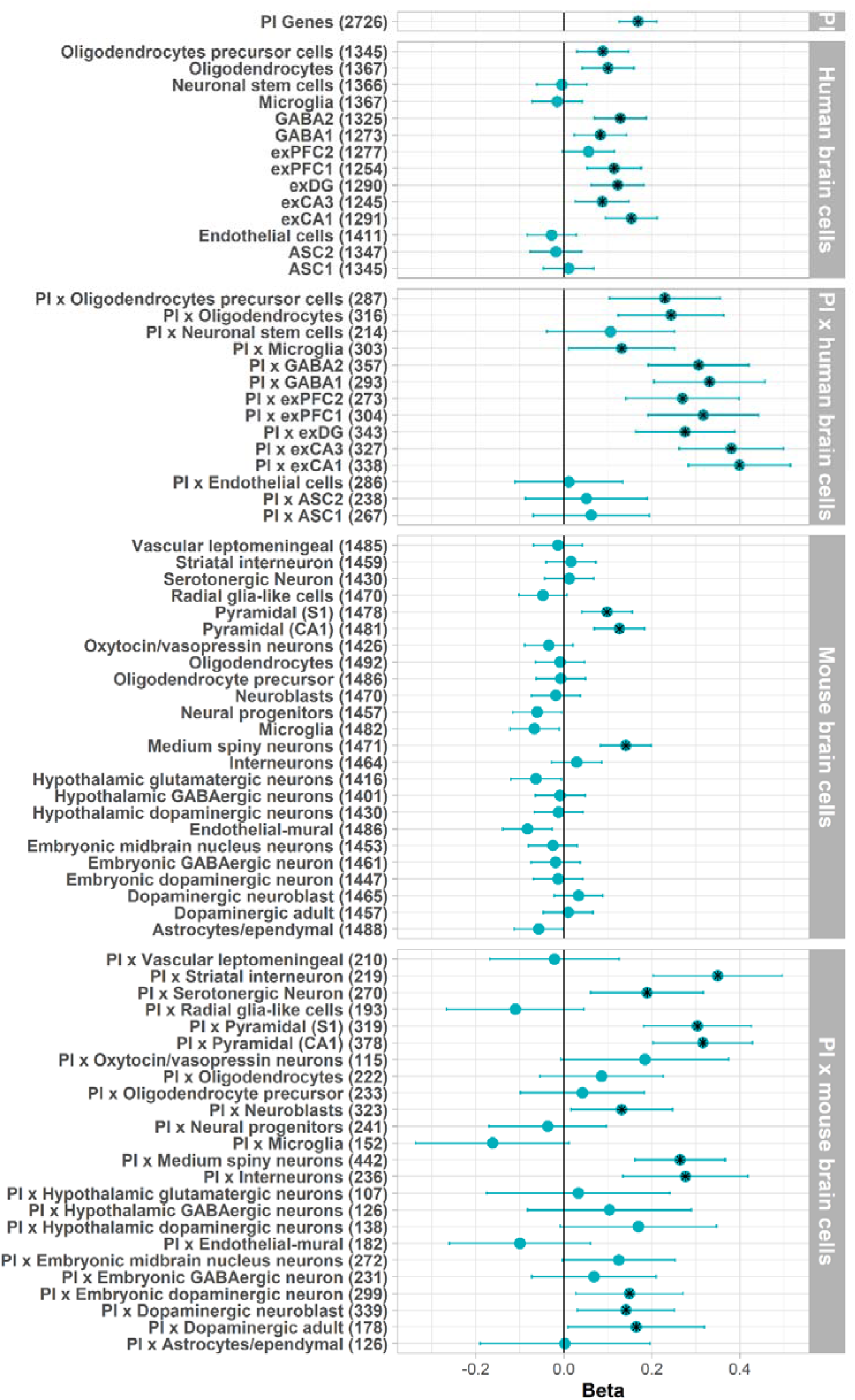
Common variant enrichment in PI and brain cell gene sets. Black stars denote significant gene-sets after multiple testing correction. ASC=astrocytes, exCA1/exCA3=pyramidal neurons from the Hippocampal *Cornu Ammonis* regions, exDG=granule neurons from the Hippocampal dentate gyrus region, exPFC1/exPFC2=pyramidal neurons from the prefrontal cortex, GABA1/GABA2=GABAergic interneurons. Although not included in the figure, the synaptic gene-sets were included in multiple testing correction.

We deviated from the pre-registration outlined by omitting parallel analyses for autism spectrum disorder. As analyses of autism exome sequence data unexpectedly required considerably more processing than the schizophrenia dataset, publication of the current valuable results for schizophrenia would be unreasonably delayed.

## RESULTS

### Common variants

In the European sample, schizophrenia-associated common variants were strongly enriched in PI genes, and genes highly expressed in human brain cells. Enrichment in genes expressed in mouse brain cells was also present across the different cell types, but not to the same extent as the human brain cells. Across brain cell types, we observed higher association betas at the intersection of PI and brain cell expressed genes suggesting stronger enrichment, although confidence intervals overlapped (Figure 1). Wider standard errors in these analyses likely reflect the loss of power due to smaller gene-sets. For human brain cell types enrichment was strongest for genes expressed in excitatory neurons from the hippocampus (pyramidal and granule) and prefrontal cortex, GABAergic interneurons and oligodendrocyte cells. In mouse brain cells enrichment was strongest in pyramidal cortical and hippocampal neurons, as well as medium spiny neurons (a type of GABAergic neuron). We observed more significantly enriched gene-sets in the PI and mouse brain intersection gene-sets, additionally implicating dopaminergic, and serotonergic neurons and interneurons. Finally, we observed significant enrichment in genes associated with synaptic processes. Common variants were most enriched in postsynaptic cellular components and biological processes. The standard errors for these associations were wide, likely due to the lower number of genes per gene-set (Supplementary Figure 4).

In analyses of individuals of East Asian ancestry, we observed a similar pattern of results as the in the analyses of European individuals, in that there was stronger enrichment at the intersection of PI and brain cell expressed genes. However, we only observed significant enrichment in four gene-sets, including the PI gene-set (Supplementary Figure 5). This may have been related to power as this GWAS had a smaller sample size. Finally, we observed a moderate correlation of 0.513 between the effect sizes from the European and East Asian analyses, which increased to 0.703 when weighted by the inverse of the product of standard errors from each association estimate (weighting precise estimates more heavily than imprecise estimates) (Supplementary Figure 6).

#### Conditional analyses

Association estimates from analyses where brain cell and intersection gene-set enrichment were compared to all other genes were similar to estimates where they were conditioned on only brain-expressed genes. We observed some attenuation when conditioning on the top 50%, 20% and 10% expressed genes (Supplementary Figure 7).

After conditional analyses of the significant intersection gene-sets, majority remained significant when conditioned on either the corresponding brain cell gene-set or PI genes, suggesting that the intersection associations are largely not confounded by the PI or brain cell gene-sets. When conditioned on both, enrichment remained significant in excitatory neurons from the hippocampus and GABAergic interneurons from human brain cells, and striatal interneurons from mouse brain cells, suggesting that the combination of both gene properties (brain cell + PI) is responsible for the enrichment of effects on schizophrenia in these genes (Supplementary Tables 2 and 3).

### (Ultra-)rare variants

After QC, 10,591 individuals (4625 cases and 5966 controls) were included in the ultra-rare variants analyses, while 10,554 individuals (4604 cases and 5950 controls) were included in the rare variant analyses. The sample sizes varied due to analysis-specific QC filters. We tested the association between the burden of rare (AF < 0.1%) or ultra-rare PTVs and schizophrenia diagnosis. Ultra-rare variants were singleton variants observed in 1 out of 188,023 individuals (our sample + gnomAD + DiscovEHR). Schizophrenia cases had a significantly higher burden of both ultra-rare (β = 0.079, SE = 0.019, *P* = 1.83 × 10^−5^) and rare (β = 0.029, SE = 0.008, *P* = 3.15 × 10^−4^) PTVs compared to controls. As a negative control, burden scores for synonymous variants were computed, which were not significantly different between cases and controls for ultra-rare (β = 0.018, SE = 0.010, *P* = 0.075) or rare (β = 0.0019, SE = 0.002, *P* = 0.445) variants. When associations were adjusted for each individual’s exome-wide (ultra-)rare variant burden, similar results were observed. Schizophrenia cases had a significantly higher burden of both ultra-rare (β = 0.066, SE = 0.019, *P* = 5.71 × 10^−4^) and rare (β = 0.024, SE = 0.008, *P* = 0.0024) PTVs compared to controls, while synonymous ultra-rare (β = -0.001, SE = 0.012, *P* = 0.926) and rare (β = -0.005, SE = 0.002, *P* = 0.083) variants remained unassociated with schizophrenia.

As rare variant analyses are defined as those with AF < 0.1%, they include singletons and thus have correlated results. Following reviewer comments we subsequently respecified rare variants as those with AF < 0.1% and excluding singletons, and tested their association with schizophrenia. This remains our reference definition of rare variants going forward. We observed no significant associations with schizophrenia case status for PTVs (β = 0.017, SE = 0.009, *P* = 0.051; adjusted for exome-wide RVs, β = 0.014, SE = 0.009, *P* = 0.110) or synonymous variants (β = 5.52 × 10^−5^, SE = 2.59 × 10^−3^, *P* = 0.983; adjusted for exome-wide RVs, β = -0.006, SE = 0.003, *P* = 0.092).

Next, we assessed (ultra-)rare variant enrichment in each gene-set using logistic regression, testing the association between the burden of (ultra-)rare variants and schizophrenia diagnosis. Rare variants (AF <0.1% + excluding singletons) were not significantly enriched in any gene-sets tested. Similar to common variants, enrichment of ultra-rare PTVs was greater at the intersection of PI genes and genes expressed in mouse and human brain cells (Figure 2), though in this case there was more enrichment of genes expressed in mouse brain cells compared to human brain cells. Across the brain cell types, enrichment was again strongest for genes expressed in excitatory neurons from the hippocampus and prefrontal cortex, as well as GABAergic interneurons, oligodendrocyte cells, and medium spiny neurons. We also observed enrichment in synaptic genes (Supplementary Figure 8). Correcting for exome-wide PTV counts reduced the number of significant gene-sets from 43 to 27 (Supplementary Figure 9). Complete regression results for PTVs and synonymous variants are described in Supplementary Tables 4 – 11.

**Figure 2.**
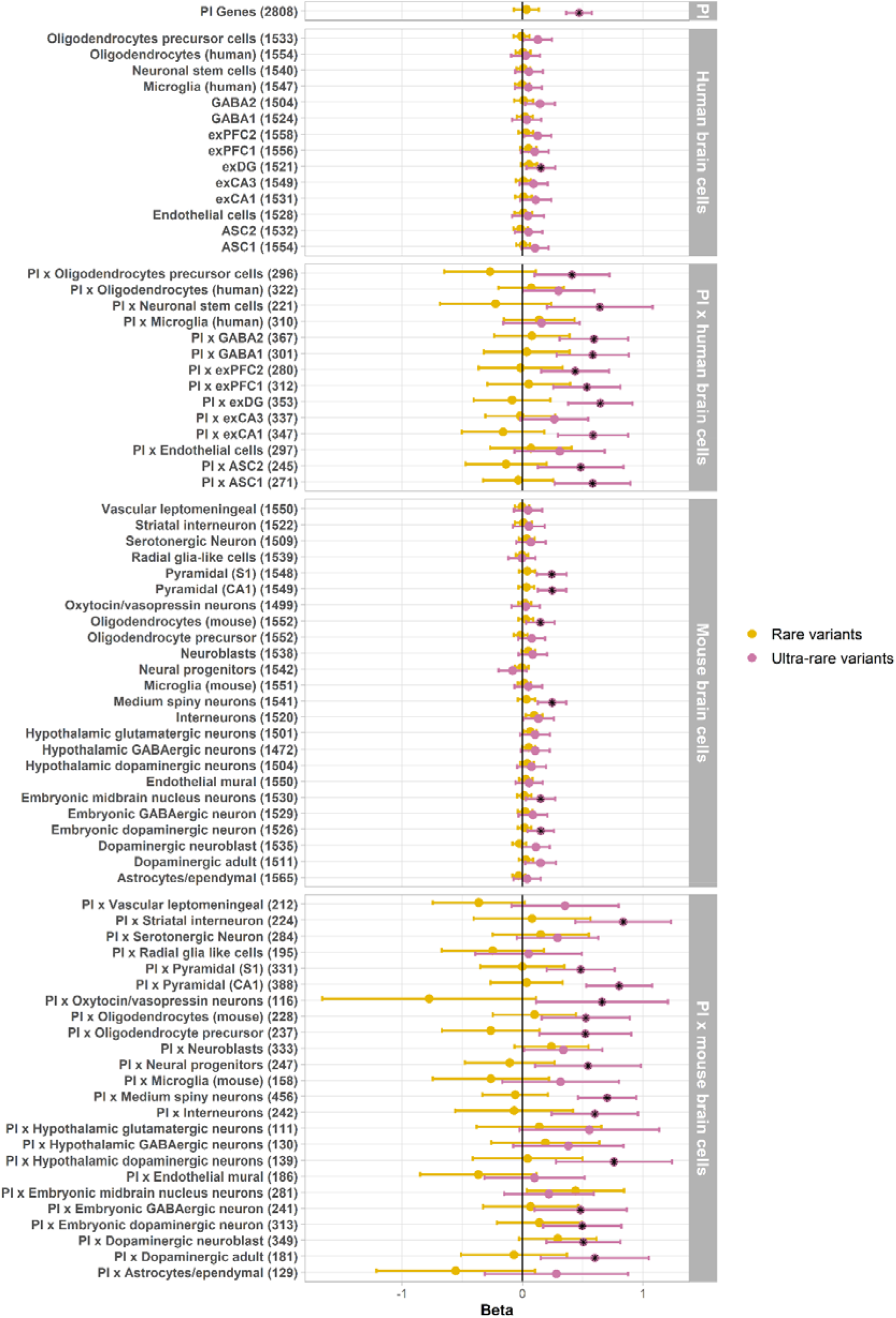
(Ultra-)rare variant enrichment in PI and brain cell gene sets. Black stars denote significant gene-sets after multiple testing correction. ASC=astrocytes, exCA1/exCA3=pyramidal neurons from the Hippocampal *Cornu Ammonis* regions, exDG=granule neurons from the Hippocampal dentate gyrus region, exPFC1/exPFC2=pyramidal neurons from the prefrontal cortex, GABA1/GABA2=GABAergic interneurons. Although not included in the figure, the synaptic gene-sets were included in multiple testing correction.

#### Conditional analyses

Significant enrichment of intersection gene-sets largely remained when corresponding brain cell gene-sets were included as covariates, however their effects were attenuated substantially when PI genes, or both were included as covariates. Although there was suggestive evidence for enrichment of PI and striatal interneuron intersection genes (Supplementary Tables 12 and 13).

### Convergence of enrichment across allele frequency spectrum

We investigated whether common and ultra-rare variants converged to similar results by rank correlating the association betas across both allele frequencies for each gene-set. We also evaluated overlapping gene-set enrichment across both allele frequencies.

We observed a correlation of 0.536 (0.707 when weighted by the inverse of the product of standard errors from each estimate) between the effect sizes of the common and ultra-rare variants, implicating PI genes, medium spiny neurons, and pyramidal neurons from the mouse brain, as well as multiple intersection gene-sets, across both allele frequencies (Figure 3). Finally, we performed a cross-ancestry comparison of the common (East Asian ancestry) and ultra-rare (European ancestry) variants. We observed a moderate correlation of 0.674 (weighted; 0.567), which suggests a shared underlying aetiology across both ancestries (Figure 4).

**Figure 3.**
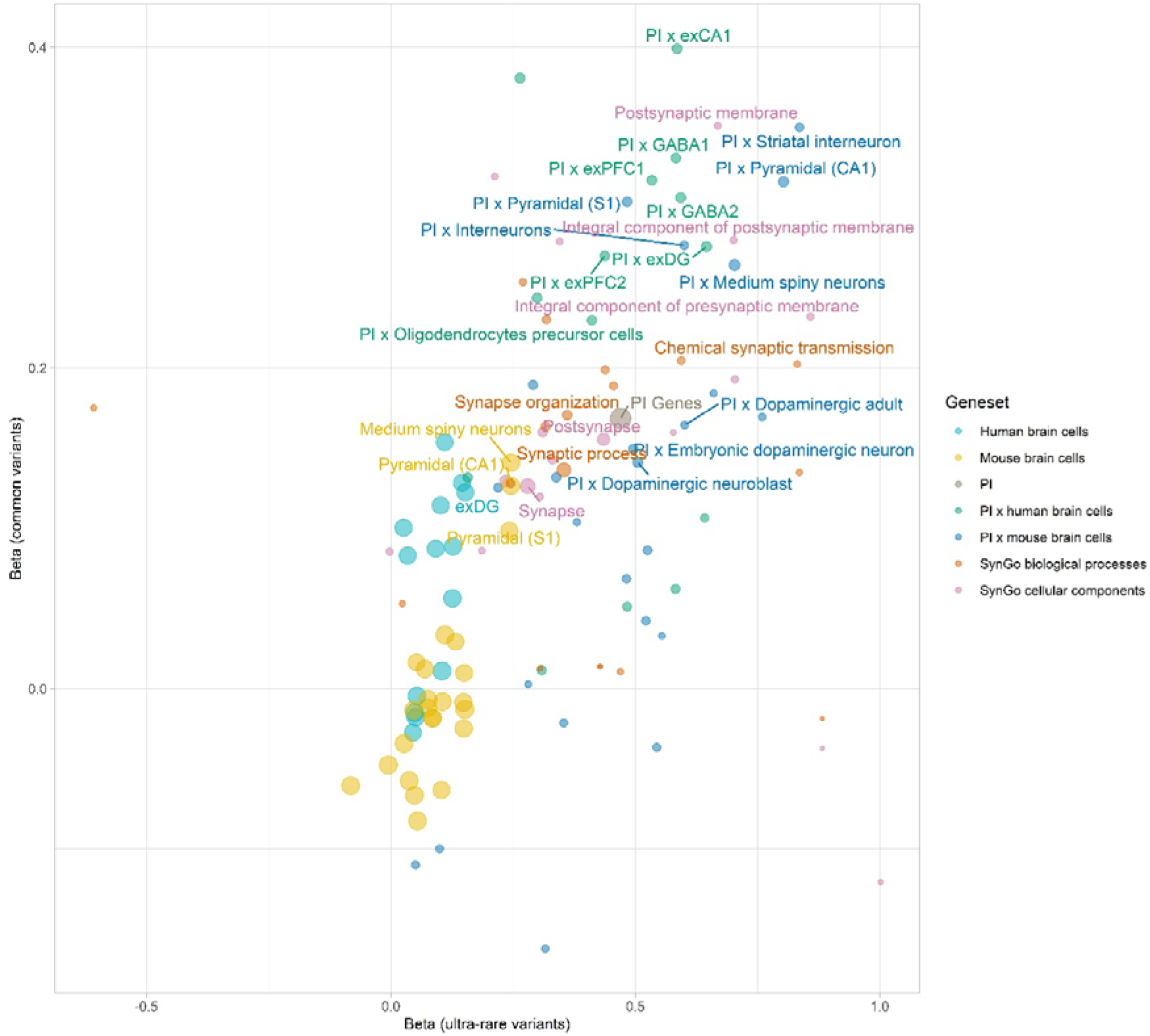
Correlation between gene set enrichment in common vs ultra-rare variants. Point sizes represent the weight assigned to each correlation estimate, obtained by calculating the inverse of the product of both standard errors. Correlation estimate is 0.536, while weighted correlation is 0.707. Labelled gene-sets are significantly enriched across both common and ultra-rare variants.

**Figure 4.**
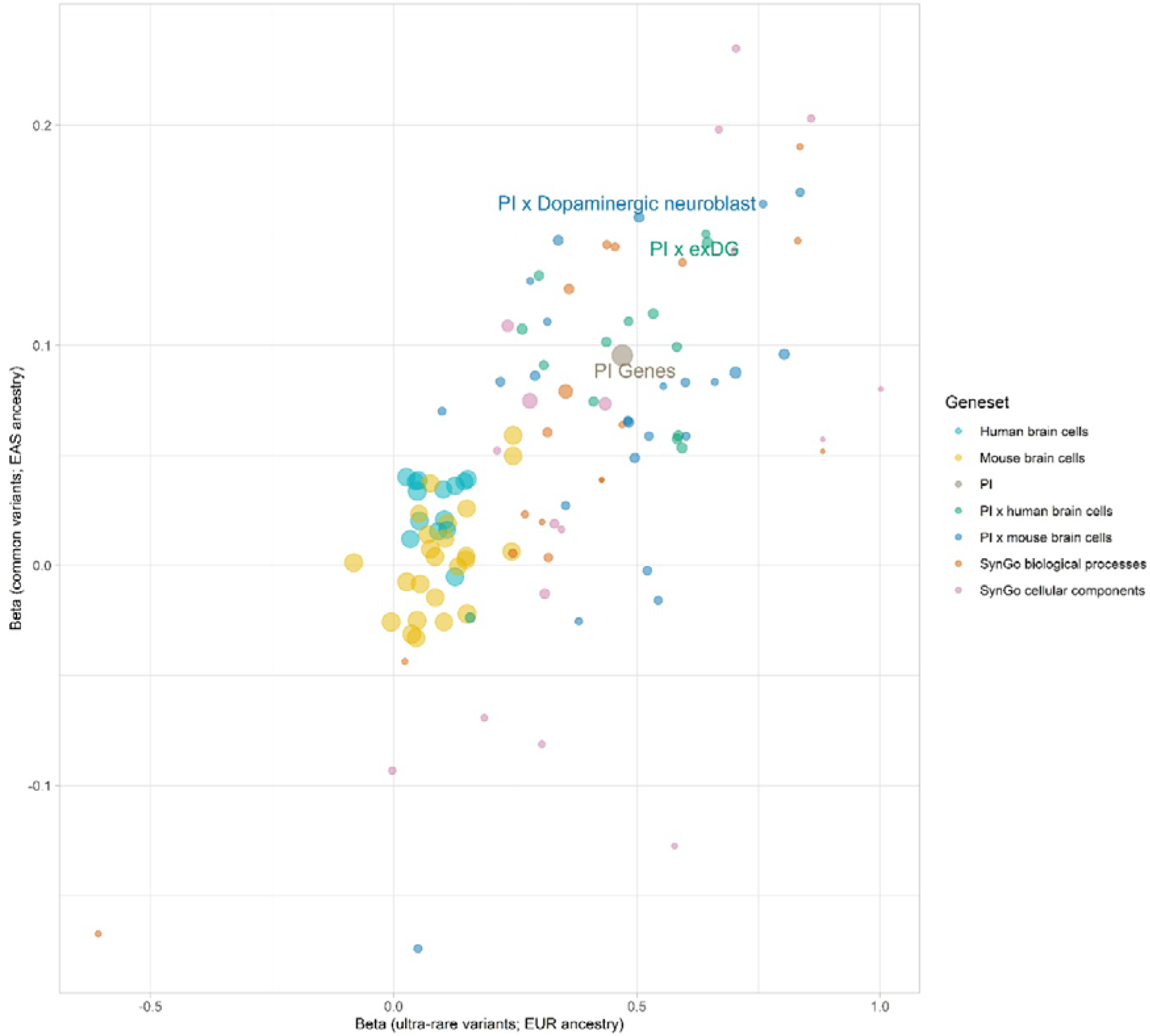
Cross-ancestry correlation between gene-set enrichment in common variants from East-Asian ancestry and ultra-rare variants from European ancestry. Point sizes represent the weight assigned to each correlation estimate, obtained by calculating the inverse of the product of both standard errors. Correlation estimate is 0.674, while weighted correlation is 0.567. Labelled genesets are significantly enriched across both common variants from individuals of East Asian ancestry and ultra-rare variants from individuals of European ancestry.

## DISCUSSION

We used GWAS and whole exome analyses of schizophrenia to investigate whether common and (ultra-)rare PTV enrichment converge to similar results in terms of what gene-sets are implicated. We observed partial convergence across the gene-sets significantly enriched for common and ultra-rare variants, in that multiple gene-sets were significantly enriched across both variant classes. Crucially, we found that after excluding singletons from their definition, no gene-sets were significantly enriched for rare variants (MAF < 0.1%). This suggests that singleton PTVs likely drive significant findings from previous analyses of rare variants. Enrichment analyses implicated mainly excitatory neurons from the prefrontal cortex and hippocampus, medium spiny neurons, and GABAergic neurons, as well as PI genes and synaptic function. Moreover, across both allele frequencies, enrichment was stronger in the gene-sets containing the intersection of brain cell types and PI genes compared to the brain cell gene-sets. Conditioning significant intersection gene-sets on both their corresponding brain cell gene-set and PI genes showed that the intersection of PI and striatal interneuron expressed genes remained significantly enriched across both common and ultra-rare variants. Finally, in cross-ancestry comparisons we observed moderate to high correlations between common variant enrichment in individuals of East Asian and European ancestries, and between common variant enrichment in individuals of East Asian ancestry and ultra-rare variant enrichment in individuals of European ancestry. This suggests a shared underlying aetiology of schizophrenia as diagnosed and operationalized across cultures.

Brain cell enrichment findings are consistent with findings from previous analyses of both common and (ultra-)rare variants associated with schizophrenia^4, 6, 18, 19^. Overlapping significant enrichment between ultra-rare and common variants provides additional evidence of some convergence in genes and biological mechanisms implicated by genetic variants across the allelic spectrum. Recent analyses showed significant enrichment of ultra-rare variants in genes implicated by schizophrenia GWAS, and that two genes implicated in rare variant analyses also showed associations in the schizophrenia GWAS^4^. Additionally, we showed that PI genes, which are likely to be under stringent selection, are implicated in both common and ultra-rare variants, while stronger enrichment in the intersection of brain cell types and PI genes suggests that PI genes are generally important, but even more so in these cell types. These gene-sets, particularly the intersection gene-sets, potentially provide a manageable set of genes and biological processes to target for follow-up analyses.

Studies of the impact of common and rare variants on complex traits have shown that they follow an inverse relationship between the variant effect sizes and population frequency, with differential selection pressures acting on variants across the allele frequency spectrum^25^. For disorders like schizophrenia which is strongly influenced by negative selection, rare variants are expected to have larger effect sizes. Indeed, our findings show that the most deleterious variants are the rarest, which can be explained by de novo mutation. Yet, hundreds of common variants explain a substantial part of schizophrenia heritability^6, 10^, and we also observed significant enrichment of common variants, which implicates similar pathways to the ultra-rare variants. A recent study has shown that common schizophrenia-associated alleles are likely maintained by background selection, a reduction of variability at neutral or nearly neutral loci as a result of negative selection against linked deleterious alleles^7, 26^. This process increases the level of linkage disequilibrium, thus alleles that are nearly neutral or mildly deleterious can be maintained due to selection against linked genes^27^. Research along both lines of enquiry are typically conducted separately, with the relevant data generally available from different consortia. As a result, directly comparable analyses of the effects of both variant classes are often difficult, even though the underlying biological/selection mechanisms that allow variants at both frequency classes to persist are likely the same/similar. Even under stringent analyses, studies including ours find overlapping signal across the allele frequency spectrum, which suggest that future studies focusing on integrating both types of analyses in the same samples are warranted.

Another factor to consider in light of our and similar findings, is how much progress the results represent with regards to biology across different diseases. Analyses of common variant cell-type enrichment support current distinctions between psychiatric disorders, and neurological disorders like Parkinson’s and Alzheimer’s disease, as they show different association patterns. Parkinson’s disease has implicated cholinergic and monoaminergic neurons, Alzheimer’s disease has implicated microglial cells, while psychiatric disorders like schizophrenia have implicated excitatory neurons^18, 28, 29^. Findings across psychiatric disorders are less clear. Analyses of individual psychiatric disorders suggest similar patterns of cell-type associations for disorders including schizophrenia, bipolar disorder, major depressive disorder (MDD), and anorexia nervosa, although enrichment was generally strongest for schizophrenia^18, 30^. However, analyses of psychiatric disorder factors have shown that common variant enrichment in excitatory and GABAergic genes from human brain cells is limited to a psychotic disorder factor (comprised of schizophrenia and bipolar disorder), with no enrichment observed in compulsive (anorexia nervosa, obsessive compulsive disorder, Tourette syndrome), neurodevelopmental (attention deficit/hyperactivity disorder, autism spectrum disorder, post-traumatic stress disorder, and MDD), or internalizing factors (post-traumatic stress disorder, MDD and anxiety disorders)^31^. Importantly, similar neuronal enrichment has been observed for non-psychiatric cognitive traits including intelligence, educational attainment, neuroticism, and body mass index (BMI), which show modest but robust genetic correlations with psychiatric disorders including schizophrenia^18, 32-34^.

These findings suggest that at the current resolution of analyses (expression differences between all cell types), common variant enrichment in genes predominantly expressed in neurons is non-specific and pervasive across various behavioural traits, although the lack of findings for psychiatric disorders may also point to differences in statistical power across the GWASs. Future analyses, for example based on more comprehensive single-cell sequencing of all neuron subtypes, could identify genes that are specific to certain developmental stages in order to seek out cell types, cell functions or developmental phases that are specific to schizophrenia. Such analyses using whole-brain developmental expression profiles have shown enrichment of schizophrenia-associated common variants in the prefrontal cortex during early midfetal development^35^. With current data it is difficult to investigate whether these similarities across various traits in common variant cell type enrichment translate to similarities in (ultra-)rare variant enrichment, and is a significant avenue for future research, although brain-expressed genes have also been found to be enriched for ultra-rare variants associated with educational attainment^36^.

Our study, as well as other results^4, 6^, further suggest that polygenicity, where very many genetic loci are implicated in a disorder like schizophrenia, complicates the search for individual risk loci in both common and ultra-rare variant analyses. As gene-set analyses are strongly correlated between common and ultra-rare variants, it is not unlikely that analyses at the level of the gene show similar correlations between common and ultra-rare variants. Strong correlations at the level of the gene may call for meta-analysis across GWAS and rare variant studies, or more subtle information integration that accounts for both the weight of evidence, as well as the regions/features/functions of the gene that are influenced by rare or common variation.

Our findings are limited by the current definitions of ultra-rare variants, and results might be subject to change if definitions change as more data becomes available. We had more power to detect effects in larger gene-sets compared to the smaller gene-sets. This was particularly evident in the intersection and synaptic gene-sets which had wide confidence intervals. Finally, our analyses were primarily focused on individuals of European and East Asian ancestry, leveraging all summary analysis available to date. While the fundamental biology of psychiatric disease, schizophrenia in particular, doesn’t vary across culture (genetic correlation between the disorder in samples of people of East Asian and European ancestry is 0.98 (SE = 0.03)^10^), the causes, presentation, and diagnosis of psychiatric disease may vary enough to have consequences for GWAS, making similar analyses in other ancestry groups crucial. A recent multi-ancestry analysis put the genetic correlation between MDD in people of European ancestry and people of East Asian ancestry at 0.41 (SE = 0.159)^37^. Analyses similar to ours typically contain limited non-European samples, with non-European samples making up 20% of the most recent schizophrenia GWAS^6^ and 26% of the most recent WES-based schizophrenia analyses^4^.

In conclusion, we show that there is at least partial overlap in the genes disrupted by both common and ultra-rare variants associated with schizophrenia suggesting involvement of the same biological mechanisms. Genes influencing neuronal processes as well as PI genes are implicated in schizophrenia aetiology across common and ultra-rare variants. Additionally, we show that singleton PTVs likely drive previous findings regarding the association between rare variants and schizophrenia. Future studies integrating information across the allele frequency spectrum might prove useful in furthering our understanding of schizophrenia aetiology.

## Data Availability

GWAS summary statistics used in the analyses are publicly available for download, with sources provided in the manuscript.
Schizophrenia exome sequenced data are available from dbGAP upon successful data application. The dbGAP study accession number is provided in the data availability section of the manuscript.  
Gene sets sources are described in the data availability section of the manuscript, and those included in the analyses are included in as supplementary materials.

https://storage.googleapis.com/gnomad-public/release/2.1.1/constraint/gnomad.v2.1.1.lof_metrics.by_gene.txt.bgz

https://gtexportal.org/home/datasets

https://static-content.springer.com/esm/art%3A10.1038%2Fnmeth.4407/MediaObjects/41592_2017_BFnmeth4407_MOESM10_ESM.xlsx

https://www.nature.com/articles/s41588-018-0129-5#Sec33

http://www.informatics.jax.org/downloads/reports/HOM_MouseHumanSequence.rpt

https://www.syngoportal.org/

## Acknowledgements

This project has received funding from the European Union’s Horizon 2020 research and innovation programme, Marie Sklodowska Curie Actions – MSCA-ITN-2016 – Innovative Training Networks under grant agreement No [721567].

W.A.A. is supported by the European Union’s Horizon 2020 research and innovation programme under the Marie Sklodowska-Curie grant agreement no. 721567, and the National Institute Of Mental Health of the National Institutes of Health under Award Number R01MH120219.

A.R.H. is supported by the Children’s Hospital Foundation and University of Queensland strategic funding.

M.B. is funded by an ERC Consolidator Grant (WELL-BEING 771057).

M.G.N is supported by the National Institute Of Mental Health of the National Institutes of Health under Award Number R01MH120219, supported by ZonMw grant: ‘Genetics as a research tool: a natural experiment to elucidate the causal effects of social mobility on health’ (pnr: 531003014) and ZonMw project: ‘Can sex- and gender-specific gene expression and epigenetics explain sex-differences in disease prevalence and etiology?’ (pnr: 849200011) from The Netherlands Organisation for Health Research and Development, a VENI grant awarded by The Dutch Research Council (NWO) (VI.Veni.191G.030), and is a Jacobs Foundation Fellow.

## DATA AVAILABILITY

- PI genes: https://storage.googleapis.com/gnomad-public/release/2.1.1/constraint/gnomad.v2.1.1.lof_metrics.by_gene.txt.bgz
- Human brain single nuclei RNA-Seq data: https://gtexportal.org/home/datasets
- Nuclei to cell type mapping: https://static-content.springer.com/esm/art%3A10.1038%2Fnmeth.4407/MediaObjects/41592_2017_BFnmeth4407_MOESM10_ESM.xlsx
- Mouse brain single cell RNA-Seq data: https://www.nature.com/articles/s41588-018-0129-5#Sec33
- Human-mouse homolog reference: http://www.informatics.jax.org/downloads/reports/HOM_MouseHumanSequence.rpt
- SynGo ontologies and annotations: https://www.syngoportal.org/
- Swedish Schizophrenia Exome Sequencing Project (**dbGaP Study Accession:** phs000473.v2.p2)

