## Supplementary Text for "Ultra-rare and common genetic variant analysis converge to implicate negative selection and neuronal processes in the aetiology of schizophrenia"

Akingbuwa et al

**Brain-expressed genes**

Data on brain-expressed genes was obtained from GTEx (Analysis V8 (dbGaP Accession phs000424.v8.p2). Of the initial 56200 genes, we selected genes that were expressed in at least 1% of brain samples in order to exclude sporadic genes and those with low expression. 30461 genes met this criteria and were further filtered to protein-coding genes that were present in the MAGMA gene location file (only overlapping genes are retained in MAGMA analyses). This resulted in 17292 brain-expressed genes to condition our brain cell and intersection gene-sets on. Subsequently, brain-expressed genes were ranked according to their median Transcripts Per Kilobase Million (TPM) values and the top 50% (8646 genes), 20% (3458 genes), and 10% (1729 genes) brain-expressed genes were further selected as gene-sets to condition on.

**Rare variant analyses**

***Quality control***

*Sample QC*: For samples where reported sex was available, we excluded individuals for which reported sex did not match genetically-derived sex. We also excluded individuals who were more than 4 median absolute deviations (MAD) from the ethnicity specific median for metrics including number of insertions, deletions, SNPs, insertion/deletion ratio, heterozygous/homozygous variants ratio, and transition/transversion (Ti/Tv) ratio. Finally, we evaluated sample relatedness between pairs of samples using identity by descent. We used a π̂ threshold of 0.2 to identify related individuals and removed one individual for each pair of first-degree relatives.

*Variant QC*: Multi-allelic variants were split prior to QC. For genotype QC we applied several filters: we removed genotypes with a depth less than 10. Additionally, for homozygous reference calls, we removed genotypes with quality less than 20; for homozygous variant calls, we filtered genotypes with less than 80% of the read depth supporting the alternate allele or with a Phred-scaled likelihood (PL) of being homozygous reference less than 20; and for heterozygous calls, we filtered genotypes with less than 80% of the read depth supporting either the reference or alternate allele, with a PL of being homozygous reference less than 20, or with less than 20% of the read depth supporting the alternate allele (i.e., an allele balance less than 0.25. Subsequently, we removed variants that were invariant after genotype filtering, as well as those that failed Variant Quality Score Recalibration (VQSR) as they are likely to be false positives. Finally, we excluded variants with Hardy-Weinberg equilibrium p-value less than 10^-6^, call rate less than 0.80, and those where the difference between case and control call rate was greater than 0.02. We also included only variants on autosomes.

*Principal component analyses:* We ran PCA analyses using largely independent high quality common variants. These were defined as having allele frequency between 0.05 and 0.95, call rate greater than 0.98 and LD-pruned to r2=0.2. We computed PCs on unrelated samples, leveraging 1000 genomes samples with known ancestry to train a random forests classifier using the PCs as features. Analyses were restricted to European samples i.e. those with greater than 0.95 probability of being European according to the random forest classifier. 201 samples from other populations including African, Ad Mixed American, East Asian and South Asian were excluded as they were too small to consider separately. We ran further PCA on strictly defined Europeans and included the first 10 PCs as covariates in association analyses.

***Variant annotations and class assignment***

We used VEP version 100^1^, defining variants either as protein-truncating (PTV), damaging missense, synonymous or non-coding.

PTVs were variants where the canonical transcript was annotated as "transcript_ablation", "splice_acceptor_variant", "splice_donor_variant", "stop_gained", or "frameshift_variant".

Damaging missense variants were annotated as "stop_lost", "start_lost", "transcript_amplification", "inframe_insertion", "inframe_deletion", "missense_variant", "protein_altering_variant", "splice_region_variant", and classified as damaging by SIFT, PolyPhen-2- HDIV, PolyPhen-2- HVAR, LRT, Mutation Taster, Mutation Assessor, and PROVEAN algorithms as ascertained from dbNSFP 3.5a.

Synonymous variants were annotated as "incomplete_terminal_codon_variant", "stop_retained_variant", or "synonymous_variant", while non-coding variants were annotated as "coding_sequence_variant", "mature_miRNA_variant", "5_prime_UTR_variant", "3_prime_UTR_variant", "non_coding_transcript_exon_variant", "intron_variant", "NMD_transcript_variant", "non_coding_transcript_variant", "upstream_gene_variant", "downstream_gene_variant", "TFBS_ablation", "TFBS_amplification", "TF_binding_site_variant", "regulatory_region_ablation", "regulatory_region_amplification", "feature_elongation", "regulatory_region_variant", "feature_truncation", or "intergenic_variant".

***Rare and ultra-rare variants definitions***

Ultra-rare variants were defined as those that were observed once in our dataset, but were absent in other publicly available WES datasets which excluded our sample, including gnomAD^2^ and DiscovEHR^3^. To account for the fact that our sample is included in gnomAD, we specified ultra-rare variants as those that were observed once in our data, but were absent in gnomAD or observed once. The rationale being that variants that are singletons in our sample with allele counts of one in gnomAD, are members of our sample and therefore can be retained as singletons. We defined rare variants as those with a minor allele frequency < 0.1% in our data set excluding ultra-rare variants. For each individual we computed separate burden scores (sum) of all (ultra-)rare variants, counting the number of variants per gene-set across both allele frequencies.

**Population stratification**

As a subset of the Swedish dataset are known to have Finnish or Northern Swedish ancestry with the potential to confound results, we performed gene-set association analyses on subsets of the data which further excluded European outliers. European outliers were determined based on the 1^st^ and 2^nd^ principal components of the Swedish sample projected on 1000 genomes data. Excluded samples are outlined in Supplementary Figure 1 below, while association betas are plotted for the different exclusionary sample subsets in Supplementary Figures 2 and 3. Regression betas from the total sample were highly correlated with betas from the different subsamples, suggesting that inclusion of different subsets did not extensively change the association statistics for the (ultra-)rare variant analyses.

**Supplementary Figure 1:** Exclusionary lines for determining possible European outliers

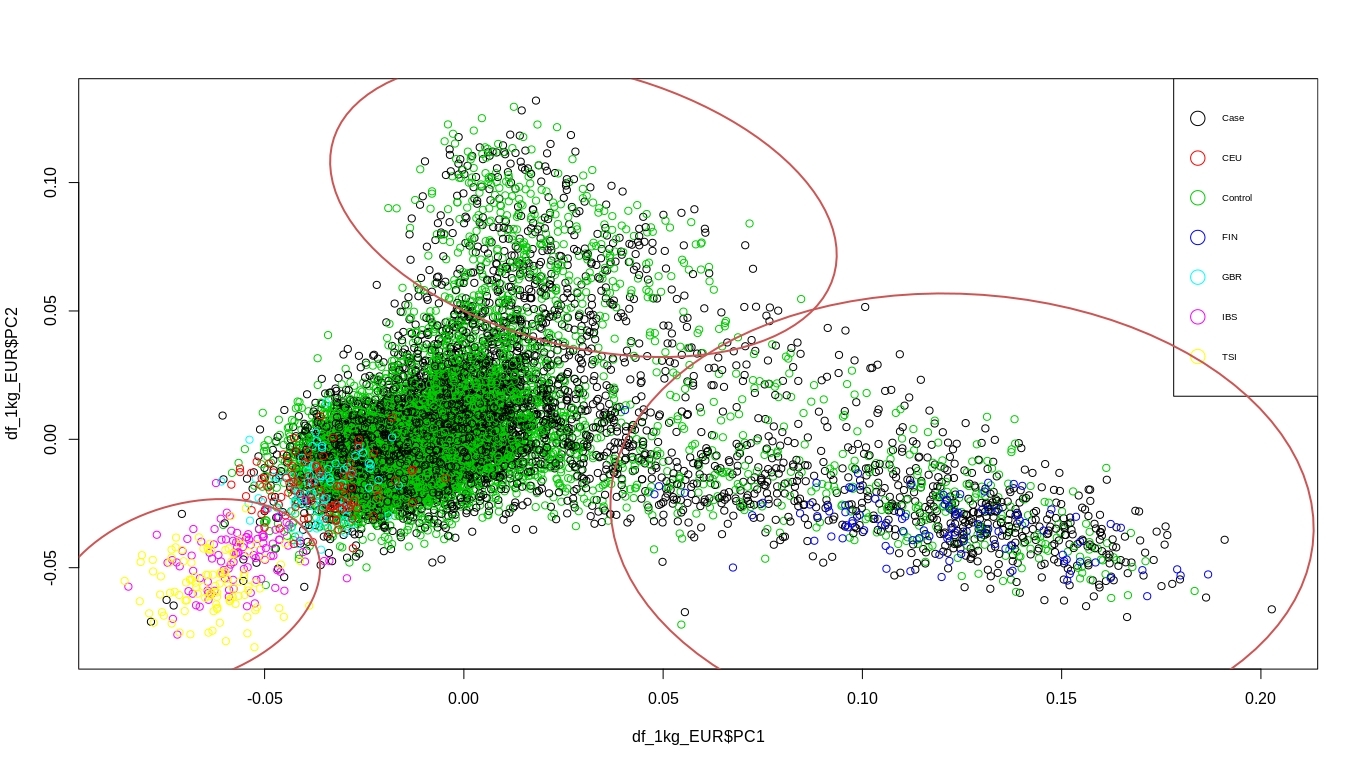

Other European

Finnish

Northern-Swedish

**Supplementary Figure 2:** Ultra-rare variant regression betas for sample subsets

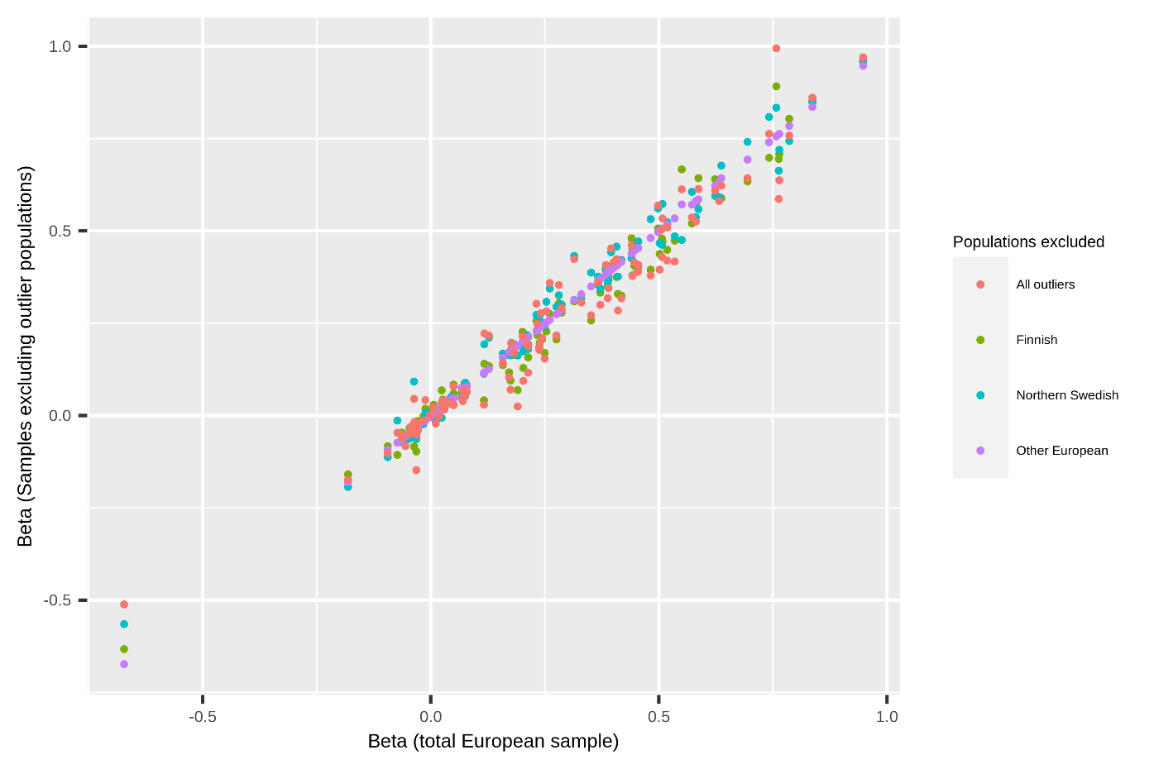

**Supplementary Figure 3:** Rare variant regression betas for sample subsets

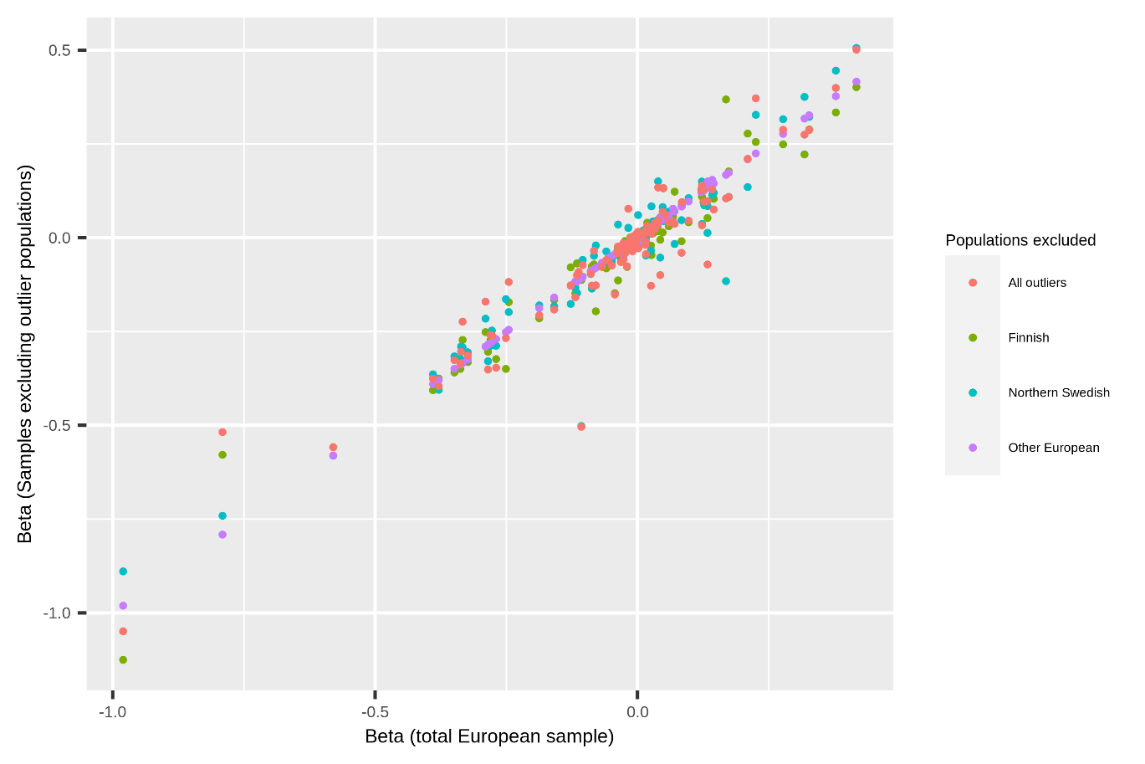

**Supplementary Figure 4**: Synaptic enrichment by schizophrenia-associated common variants

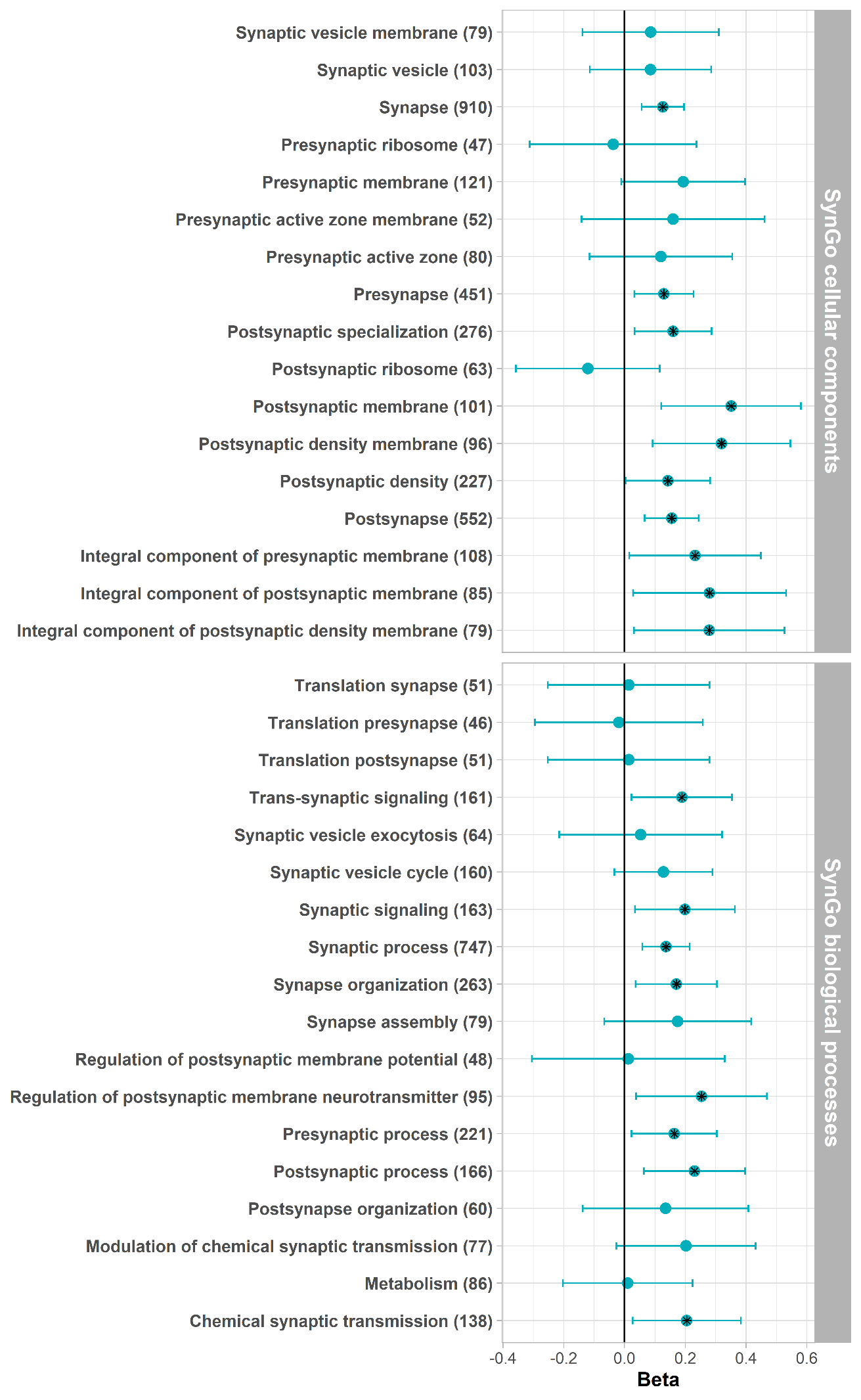

**Supplementary Figure 5**: Enrichment by schizophrenia-associated common variants in individuals of East-Asian Ancestry

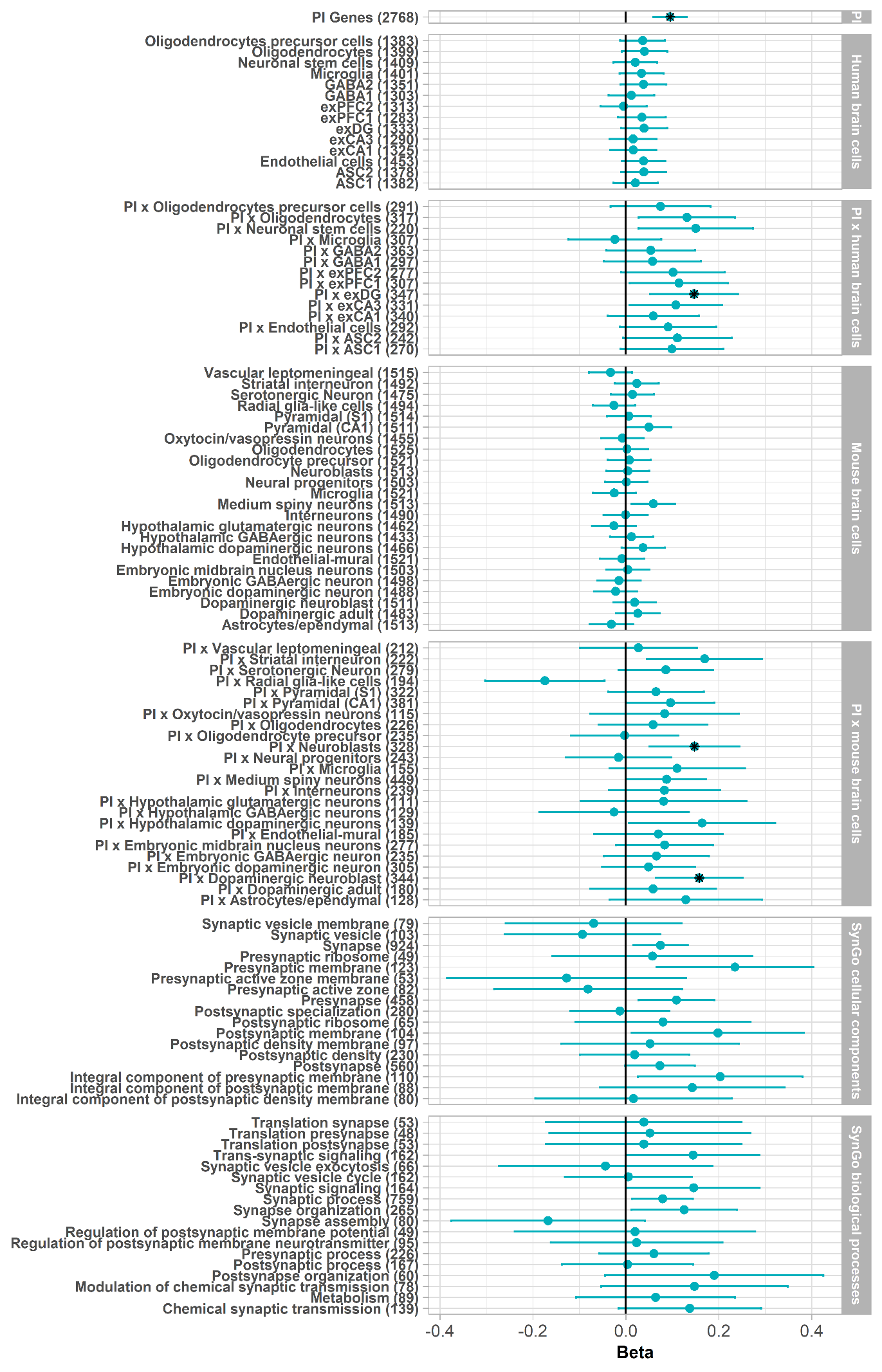

**Supplementary Figure 6**:

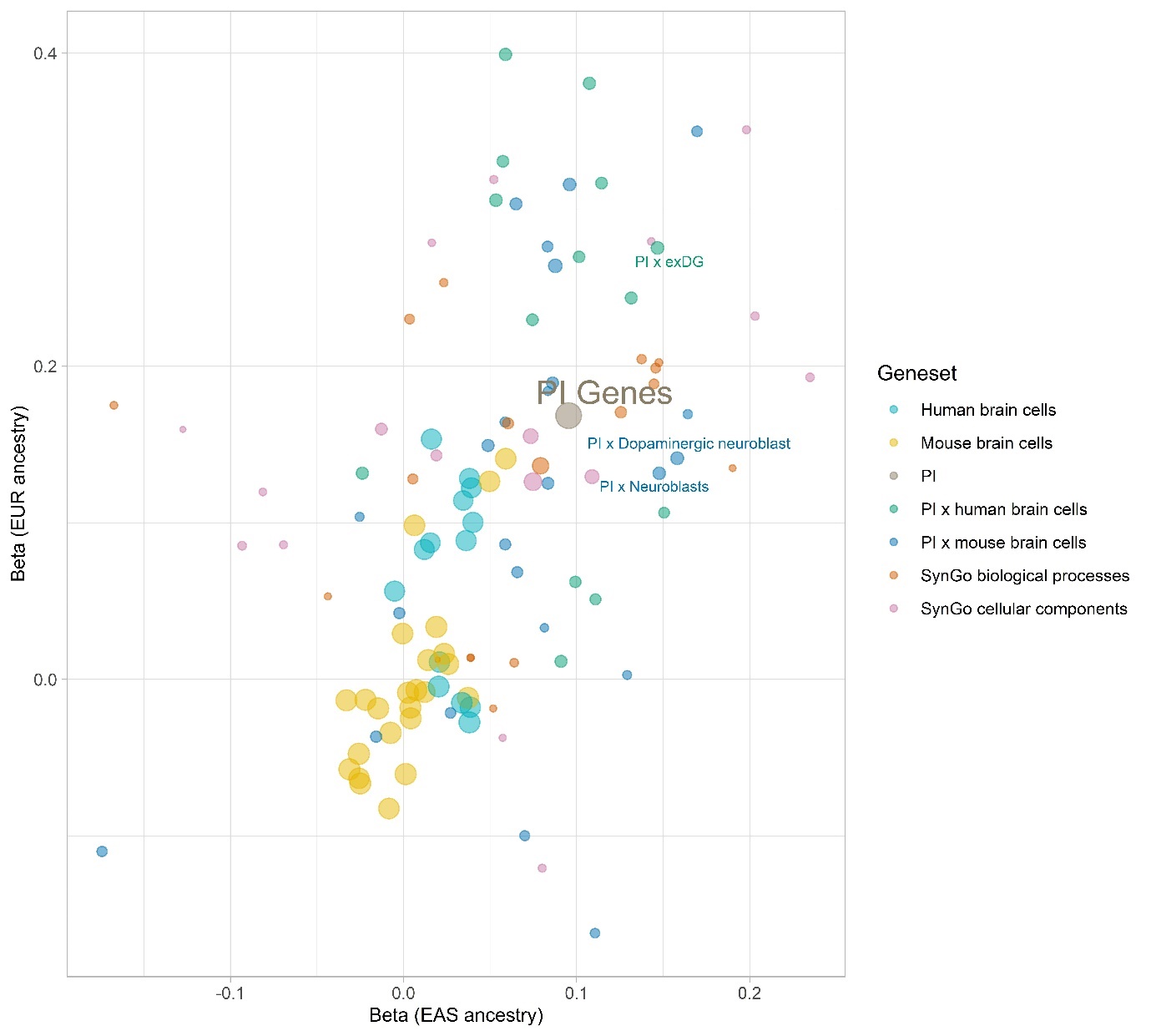

Correlation between common variants gene-set enrichment in European vs East Asian ancestry. Labelled gene-sets are significantly enriched across both common and rare variants**.** Point sizes represent the weight assigned to each correlation estimate, obtained by calculating the inverse of the product of both standard errors.

**Supplementary Figure 7**: Enrichment in brain cell gene-sets conditional on only brain expressed genes

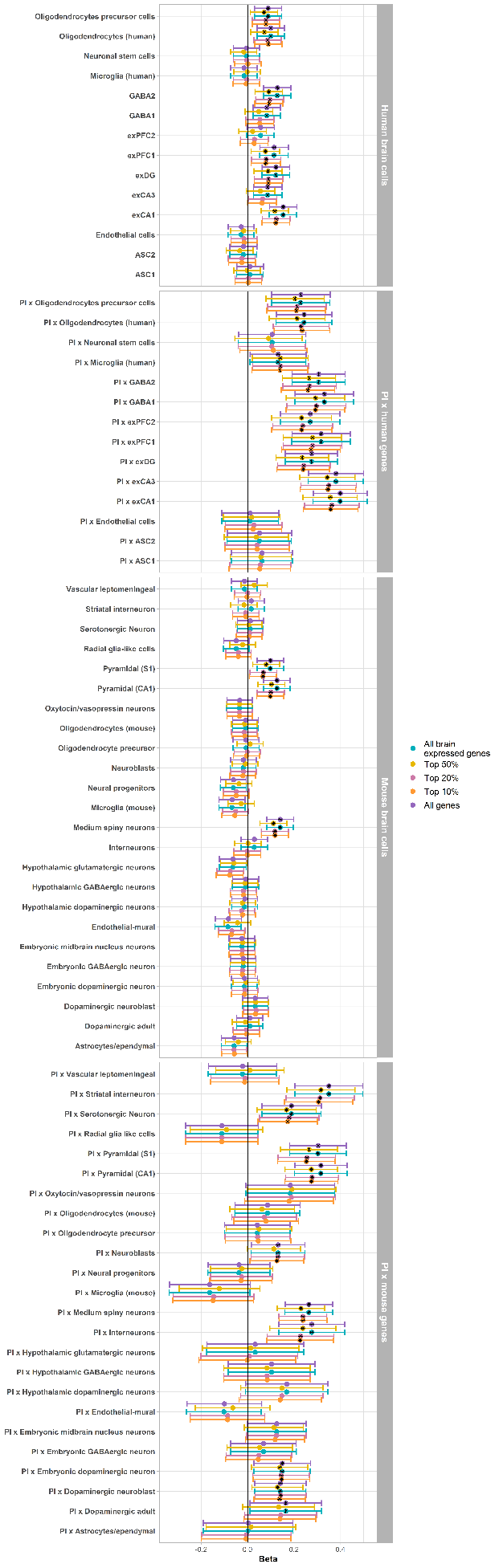

**Supplementary Figure 8**: Synaptic enrichment by schizophrenia-associated (ultra-)rare variants

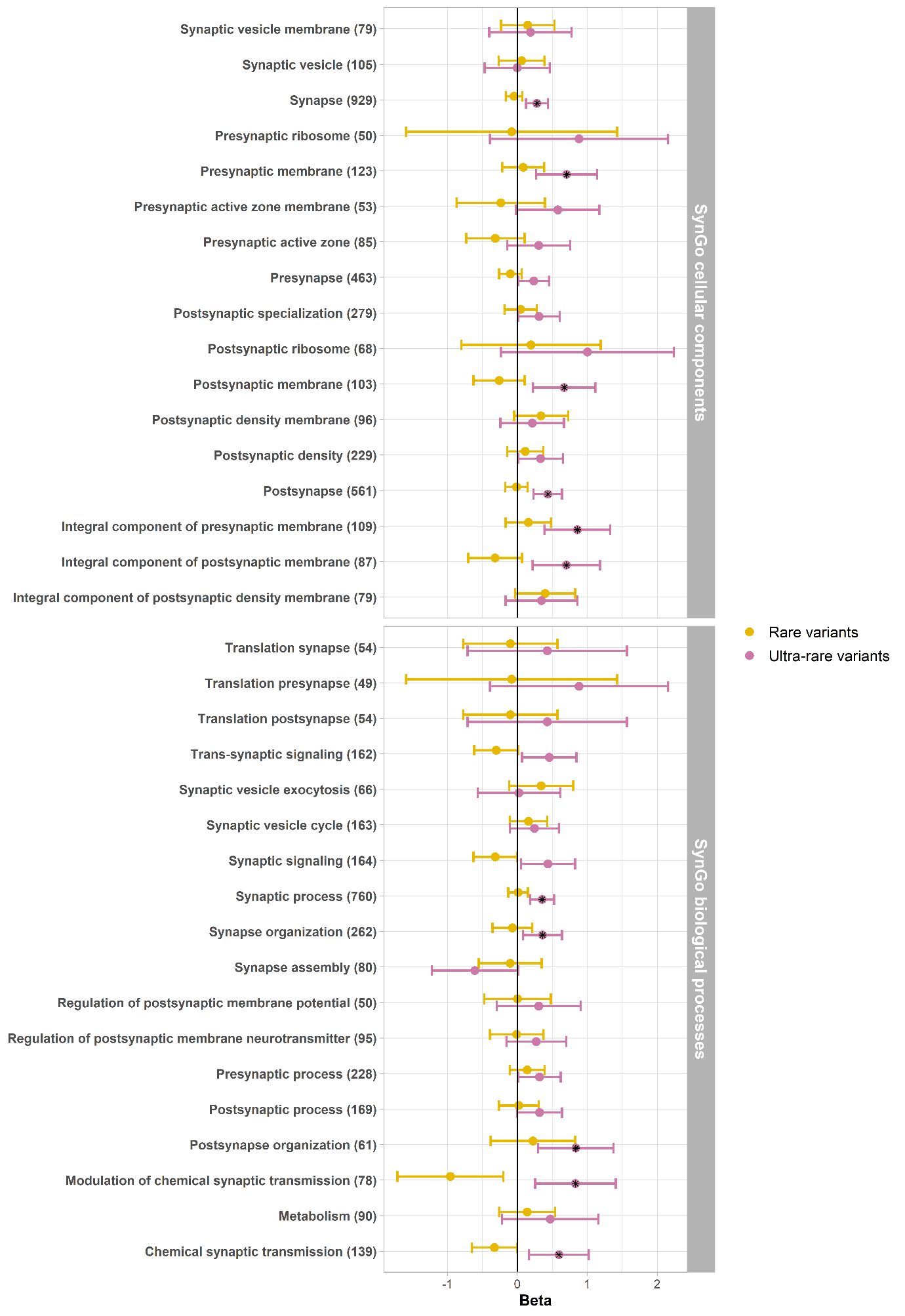

**Supplementary Figure 9**: (Ultra-)rare variant enrichment of PTVs after adjusting for exome-wide PTV burden

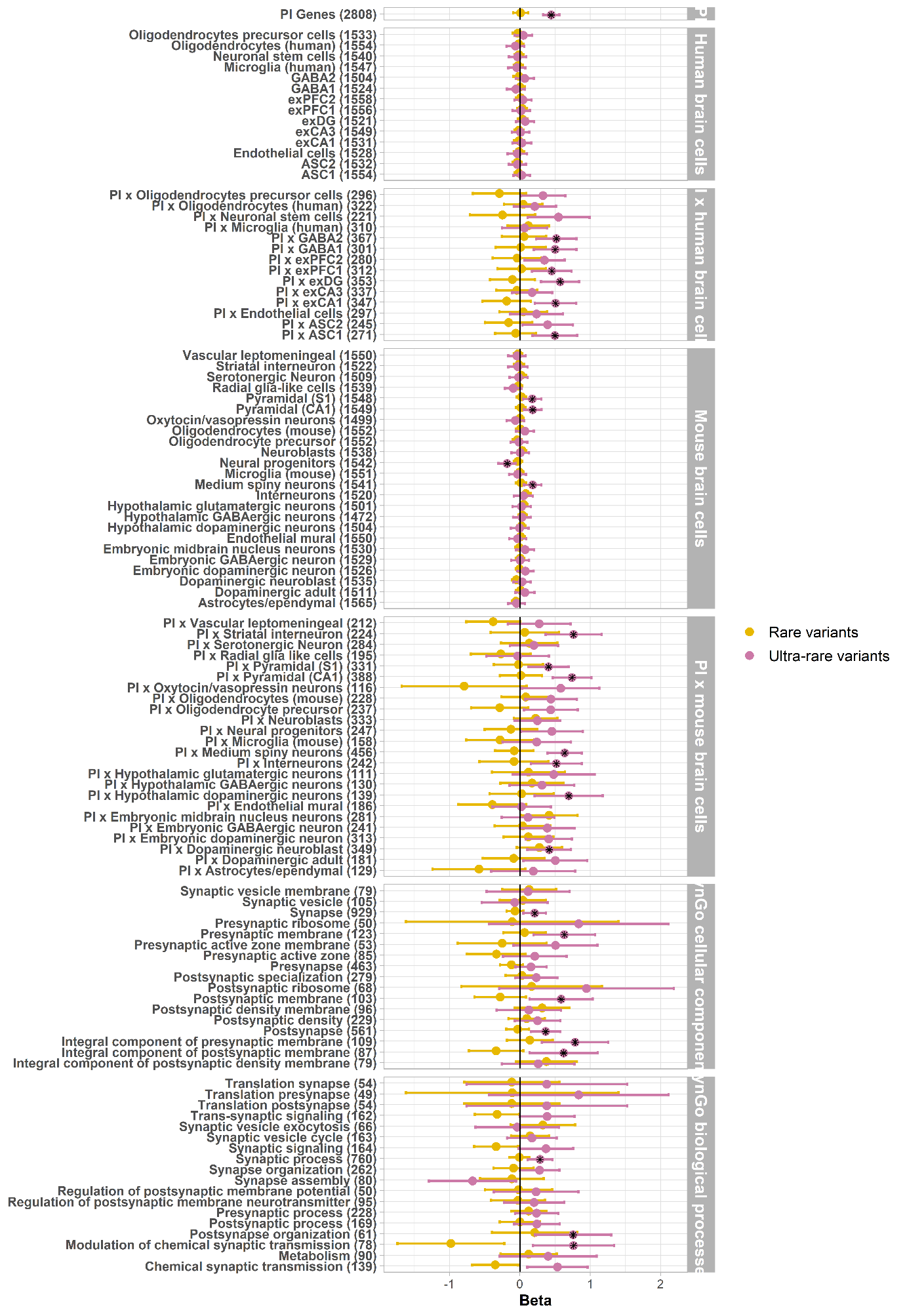
